# SARS-CoV-2 Omicron Neutralization After Heterologous Vaccine Boosting

**DOI:** 10.1101/2022.01.13.22268861

**Authors:** Kirsten E. Lyke, Robert L. Atmar, Clara Dominguez Islas, Christine M. Posavad, Daniel Szydlo, Rahul PaulChourdhury, Meagan E. Deming, Amanda Eaton, Lisa A. Jackson, Angela R. Branche, Hana M. El Sahly, Christina A. Rostad, Judith M. Martin, Christine Johnston, Richard E. Rupp, Mark J. Mulligan, Rebecca C. Brady, Robert W. Frenck, Martín Bäcker, Angelica C. Kottkamp, Tara M. Babu, Kumaravel Rajakumar, Srilatha Edupuganti, David Dobrzynski, Rhea N. Coler, Janet I. Archer, Sonja Crandon, Jillian A. Zemanek, Elizabeth R. Brown, Kathleen M. Neuzil, David S. Stephens, Diane J. Post, Seema U. Nayak, Paul C. Roberts, John H. Beigel, David Montefiori, the DMID 21-0012 Study Group

## Abstract

As part of an ongoing study assessing homologous and heterologous booster vaccines, following primary EUA series, we assessed neutralization of D614G and Omicron variants prior to and 28 days after boost. Subset analysis was done in six combinations (N = 10/group): four homologous primary-booster combinations included mRNA-1273 two-dose priming followed by boosting with 100-μg or 50-μg mRNA-1273, Ad26.COV2.S single-dose priming followed by Ad26.COV2.S booster and BNT162b2 two-dose priming followed by BNT162b2 boosting; and two heterologous primary-booster combinations: BNT162b2 followed by Ad26.COV2.S and Ad26.COV2.S followed by BNT162b2. Neutralizing antibody (Nab) titers to D614G on the day of boost (baseline) were detected in 85-100% of participants, with geometric mean titers (GMT) of 71-343 in participants who received an mRNA vaccine series versus GMTs of 35-41 in participants primed with Ad26.OV2.S. Baseline NAb titers to Omicron were detected in 50-90% of participants who received an mRNA vaccine series (GMT range 12.8-24.5) versus 20-25% among participants primed with Ad26.COV2.S. The booster dose increased the neutralizing GMT in most combinations to above 1000 for D614G and above 250 for Omicron by Day 29. Homologous prime-boost Ad26.COV2.S had the lowest NAb on Day 29 (D614G GMT 128 and Omicron GMT 45). Results were similar between age groups. Most homologous and heterologous boost combinations examined will increase humoral immunity to the Omicron variant.

---

The Severe Acute Respiratory Syndrome Coronavirus 2 (SARS-CoV-2) Omicron (B.1.1.529) variant of concern, first reported to the World Health Organization on November 24, 2021, has become the dominant circulating strain in the United States.^1^ The large number of mutations, including 15 within the spike protein’s receptor binding domain, are associated with increased transmissibility, escape from therapeutic antibodies, and partial resistance to natural- and vaccine-induced immunity, with increasing breakthrough infections.^2.3^ As part of an ongoing study assessing homologous and heterologous booster vaccines,^4^ we assessed neutralization of D614G and Omicron variants.

Six combinations were assessed: four homologous primary-booster combinations included mRNA-1273 (Moderna, two doses of 100-μg) followed by boosting with 100-μg or 50-μg mRNA-1273, Ad26.COV2.S (Janssen, one dose of 5×10^10^ virus particles) followed by the same dose of Ad25.COV2.S and BNT162b2 (Pfizer-BioNTech, two doses of 30-μg) followed by 30-μg of BNT162b2; and two heterologous primary-booster combinations: BNT162b2 followed by Ad26.COV2.S and Ad26.COV2.S followed by BNT162b2, with booster intervals ranging from 3-6.5 months. Samples from a randomly selected subset of 20 participants per group (10 participants 18-55 years old and 10 participants ≥56 years) were assayed. We determined 50% serum neutralizing antibody (NAb) titers using a lentivirus-based pseudovirus assay^5^ against the D614G reference standard (reported previously^4^ for all groups but the 50-μg mRNA-1273 homologous boost group) and Omicron spike variants from Day 1 (pre-boost) and Day 29 post-boost.^4^

NAb titers to D614G on the day of boost (Day 1, baseline) were detected in 85-100% of participants, with geometric mean titers (GMT) of 71-343 in participants who received an EUA mRNA vaccine series versus GMT 35-41 in participants primed with Ad26.OV2.S (supplemental material). Baseline NAb titers to Omicron were detected in 50-90% of participants who received an EUA mRNA vaccine series (GMT range 12.8-24.5) versus 20-25% among participants primed with Ad26.COV2.S (GMT range 7.2-7.6). Titers were 6.0-14.0-fold lower relative to D614G. The booster dose increased the neutralizing GMT in most combinations to above 1000 for D614G and above 250 for Omicron (Figure). Homologous prime-boost Ad26.COV2.S had the lowest NAb on Day 29 (D614G GMT 128 and Omicron GMT 45). After boost, the GMTs were 2.3-7.5-fold lower for Omicron as compared to D614G. Results were similar between age groups.

**Figure 1.**
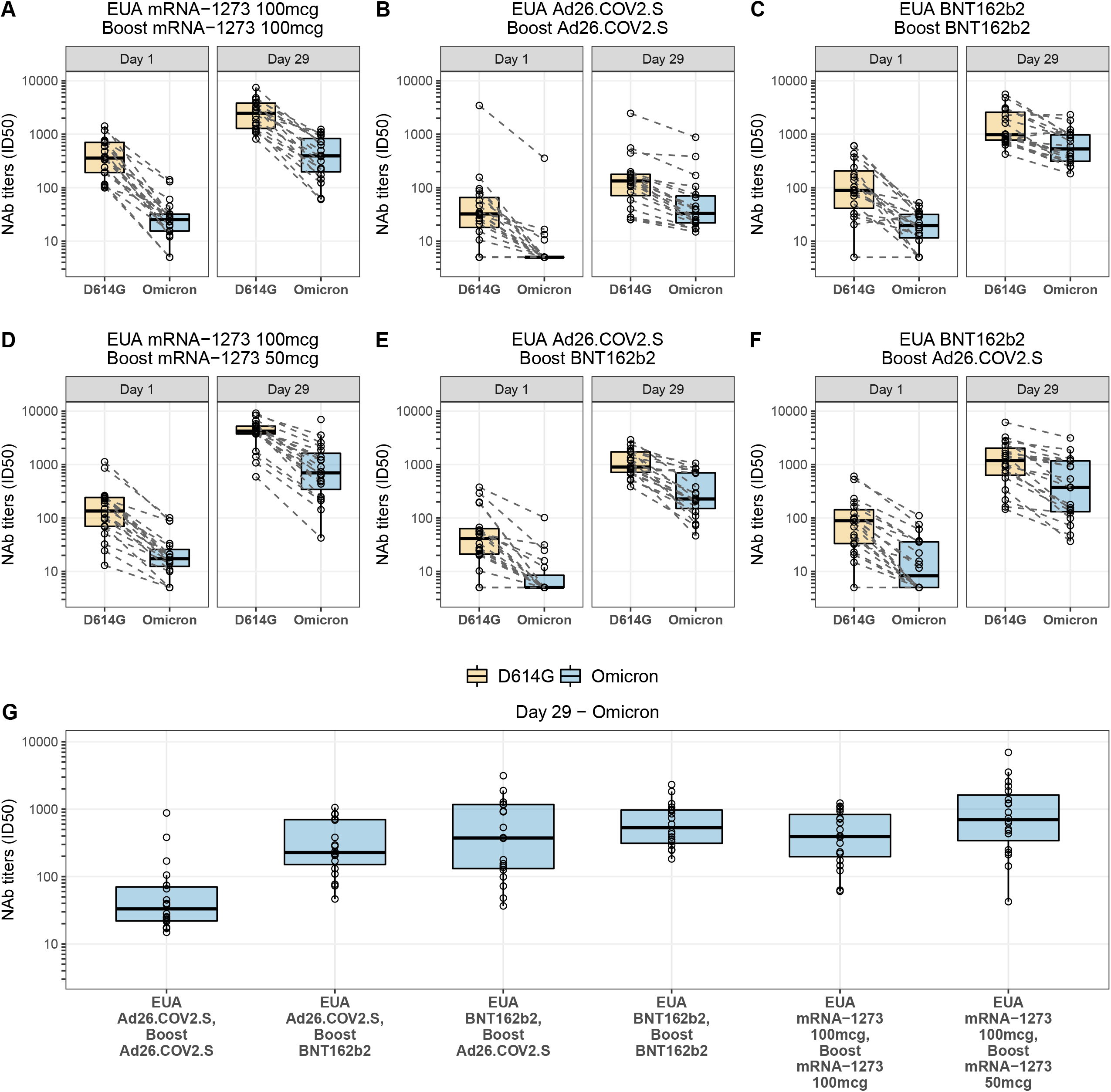
Pseudovirus neutralization expressed as 50% inhibitory dilution (ID50) to D614G and Omicron at Day 1 (pre-booster) and Day 29 for the combinations: A) mRNA-1273 EUA vaccine boosted with mRNA-1273 100-μg; B) Ad26.COV2.S EUA vaccine boosted with Ad26.COV2.S; C) BNT162b2 EUA vaccine boosted with BNT162b2; D) mRNA-1273 EUA vaccine boosted with mRNA-1273 50-μg; E) Ad26.COV2.S EUA vaccine boosted with BNT162b2; and F) BNT162b2 EUA vaccine boosted with Ad26.COV2.S. G) post-boost pseudovirus neutralization to Omicron for all combinations. Box plots represent median (horizontal line within the box) and 25th and 75th percentiles (lower and upper borders of the box), with the whiskers drawn to the value nearest to, but within, 1.5 x IQR above and below the borders of the box.

Homologous or heterologous boosts with available EUA vaccines provide post-boost titers to Omicron that have been associated with >85% protection against symptomatic SARS-CoV-2 infection with D614G and B.1.1.7 (Alpha), in most combinations tested.^5^ Homologous prime-boost Ad26.COV2-S is the exception, with post-boost NAb being lower than other combinations. All combinations tested decreased the difference in titers between D614G and Omicron after boost. The 50-ug Moderna booster achieved similar NAb titers against Omicron as the 100-ug dose, including in those ≥56 years. Although vaccine-induced nNAb correlate with protection against illness, real world effectiveness data and better understanding of the role of cell-mediated immunity and correlates of protection against severe disease are needed. Additionally, the kinetics of the NAb post-boost are unknown - a factor critical to understanding the need for additional boosters Taken together, these data support that most homologous and heterologous boost combinations will increase humoral immunity to Omicron and can be considered as a strategy to mitigate risk from this variant.

## Data Availability

All data produced in the present study are available upon reasonable request to the authors.

## Funding and Acknowledgements

The trial was sponsored and primarily funded by the Infectious Diseases Clinical Research Consortium through the National Institute for Allergy and Infectious Diseases (NIAID) of the National Institutes of Health (NIH), under award numbers UM1AI48372, UM1AI148373, UM1AI148450, UM1AI148452, UM1AI148573, UM1AI148574, UM1AI148575, UM1AI148576, UM1AI148684, UM1 AI148689 and with support from the NIAID Collaborative Influenza Vaccine Innovation Centers (CIVICs) contract 75N93019C00050 and NIH Vaccine Research Center.

We would like to acknowledge Moderna, Inc., Johnson & Johnson/Janssen, and Pfizer/BioNTech Pharmaceuticals for their collaboration, scientific input, and sharing of documents needed to implement this trial. All products were acquired through the government procurement process.

