## Supplementary Appendix for "SARS-CoV-2 Omicron Neutralization After Heterologous Vaccine Boosting"

This appendix is submitted by the authors to provide additional information about their work.

**Supplement to:** Lyke KE*, Atmar RL*, et al. Neutralizing Antibody Responses to the SARS CoV-2 Omicron Variant after Homologous or Heterologous Booster Vaccination

*Contributed equally

Supplementary Appendix to Manuscript Entitled

### Neutralizing Antibody Responses to the SARS CoV-2 Omicron Variant after Homologous or Heterologous Booster Vaccination

Mix and Match Study Group
(listed in PubMed and ordered by lead institutions and enrollment)

**University of Maryland, Institute of Human Virology and Center for Vaccine Development and Global Health, Baltimore, MD, VTEU**
Kirsten E. Lyke, M.D., Meagan Deming, M.D., Ph.D., Jennifer S. Husson, M.D., M.P.H., Angie Price, D.N.P., M.S.N., C.R.N.P.

**Baylor College of Medicine (BCM), Houston TX, Vaccine Training and Evaluation Unit (VTEU)**
Robert L. Atmar, M.D., Hana El Sahly, M.D., Jennifer A. Whitaker, M.D., and Wendy A. Keitel, M.D.

**Kaiser Permanente Washington Health Research Institute, Seattle, Washington, VTEU**
Lisa A. Jackson, MD, MPH

**University of Rochester Medical Center, Rochester, NY, VTEU**
Angela Branche, M.D., David Dobrzynski, M.D., Ann R. Falsey, M.D., Ian Shannon, MS, RN,

**The Hope Clinic of Emory University (Hope), Atlanta, GA, VTEU**
Srilatha Edupuganti, M.D., M.P.H., Daniel Graciaa, M.D., Nadine Rouphael, M.D., M.Sc.

**Emory Children's Center (ECC), Atlanta, GA, VTEU**
Christina Rostad, M.D., Evan J. Anderson, M.D., Satoshi Kamidani, M.D.

**University of Pittsburgh School of Medicine, Pittsburgh, PA, subsidiary to Vanderbilt University VTEU, Nashville, TN**
Judith M. Martin, M.D., Kumaravel Rajakumar, M.D., M.S., Gysella B. Muniz M.D., Sonika Bhatnagar, M.D., M.P.H.

**University of Washington, Seattle, WA, VTEU**
Christine Johnston, M.D., M.P.H., Tara M. Babu, M.D., M.SCI., Anna Wald, M.D., M.P.H.

**University of Texas Medical Branch (UTMB), League City, TX, subsidiary to BCM VTEU**
Richard E. Rupp, M.D., Megan Berman, M.D., Laura Porterfield, M.D., Amber Stanford, PA-C

**New York University – Bellevue Vaccine Center, New York, NY, VTEU**
Mark J. Mulligan, M.D., FIDSA, Angelica Cifuentes Kottkamp, M.D., Jennifer Lee Dong, M.D.

**New York University- Long Island Vaccine Center, Mineola, NY, subsidiary to NYU VTEU**
Martín Bäcker, MD, Steven E. Carsons, MD, Diana Badillo, MD,

**Cincinnati Children's Hospital Medical Center (CCHMC), Cincinnati, OH, VTEU**
Rebecca Brady, M.D., Robert W. Frenck, Jr., M.D., Susan Parker, R.N., Michelle Dickey, A.P.R.N.

**Seattle Children's Research Institute, Seattle, WA**
Rhea N. Coler, M.Sc., Ph.D., Sasha E. Larsen, Ph.D.

**Infectious Diseases Clinical Research Consortium (IDCRC) Laboratory Operations Unit (LOU)**
Christine M. Posavad, Ph.D., John Hural, Ph.D

**IDCRC Leadership**
Kathleen M. Neuzil, M.D., David S. Stephens, M.D.

**IDCRC Statistical and Data Science Unit (SDSU):**
Elizabeth R. Brown, Sc.D., Clara Dominguez Islas, Ph.D.

**Statistical Center for HIV/AIDS Research and Prevention (SCHARP), Seattle, WA:**
Jillian Zemanek, M.P.H., Daniel Szydlo, M.S., Rahul PaulChoudhury, M.S.,

**Division of Microbiology and Infectious Diseases (DMID), National Institute of Allergy and Infectious Diseases (NIAID), National Institutes of Health (NIH), Bethesda, MD.**
Sonja Crandon, B.S.N., Diane J. Post, Ph.D., Marina Lee, Ph.D., Seema Nayak, M.D., Paul C Roberts, Ph.D., John Beigel, M.D.

**FHI360, Durham, NC**
Janet I. Archer M.Sc.

**Duke University Laboratory – Durham, NC**
David Montefiori, Ph.D., Amanda Eaton, Ph.D.

#### Mix and Match Study Team Members

**University of Maryland, Institute of Human Virology and Center for Vaccine Development and Global Health, Baltimore, MD, VTEU**

Kirsten E. Lyke, M.D., Meagan E. Deming, M.D., Ph.D., Jennifer S. Husson, M.D., M.P.H., Angie Price, D.N.P., M.S.N., C.R.N.P., Joel Chua, M.D., Karen Kotloff, M.D., Myounghee Lee, Pharm.D., Ph.D., Lisa Anderson, R.N., B.S., Amy Nelson, R.N., M.S., Salma Sharaf, Young Chae Jessica Yoo, Lisa Langer, Pharm.D., Brenda Dorsey, R.N., B.S., M.S., CCRP., Alyson Kwon, M.S., CCRC, ACRP-PM., Sophie Harper, M.Sci., Paula Bernal, M.Sci, Ph.D., Eric Goldstein, Jeffrey Floyd, Marcelo Sztein, M.D., Ming Bell, M.D., Leslie Howe, Erika Stiles, Andrew Chi, Pharm.D., Phuong Tran Nguyen, Pharm.D., Yogitha Pazhani, Pharm.D., Christine Aggabao, Pharm.D., Jeannie Murray.

**Baylor College of Medicine (BCM), Houston TX, Vaccine Training and Evaluation Unit (VTEU)**

Robert L. Atmar, M.D.; Hana El Sahly, M.D.; Jennifer A. Whitaker, M.D.; Wendy A. Keitel, M.D.; Mary Healy, M.D.; Christine Akamine, M.D.; Pedro A. Piedra, M.D.; Chanei Henry, A.A.S.; Brandie Phillips, R.N.: Chianti Wade-Bowers, B.S.N, M.S.N.; Connie Rangel, R.N.; Logan Lee; Julia Guardado; Angelica Diaz, F.N.P.; Lisreina Toro; Tina Sierra; Janet Brown, J.D., R.Ph. : Cathy Faw, R.Ph.; Yvette Rugeley; Yolanda Rayford; Kayla Burrell; Jesus Banay; Tykel Eddy; Marinna Matta; Kathy Bosworth, B.A.

**Kaiser Permanente Washington Health Research Institute, Seattle, WA, VTEU**

Lisa A. Jackson, MD, MPH, Lee Barr, R.N., Jesse Berg, B.S., Cassandra Bryant, B.S., Roger Calvert, PA-C, Barbara Carste, M.P.H., Joe Choe, B.S., Maya Dunstan, M.S., R.N., Jana ffitch, L.P.N., Colin Fields, M.D., Lynn Gross, PA-C, Erika Kiniry, M.P.H., Bonnie Lam, PharmD, De Vona Lang, Rebecca Lau, PharmD, Stella Lee, B.A., Paula Lins, PA-C, Amy Mohelnitzky, PA-C, Marilyn Nguyen, B.S., Matthew Nguyen, M.P.H., Hallie Phillips, M.Ed. Stephanie Pimienta, B.S., Melissa Resendiz Rivas, B.A., Melissa Boothe Scheer, PA-C, Janice Suyehira, M.D

**Seattle Children's Research Institute, Seattle, WA subsidiary to Kaiser Permanente VTEU**

Rhea N. Coler, M.Sc., Ph.D., Sasha E. Larsen, Ph.D., Evan Cross, B.Sc., Tiffany Pecor, B.S., Thomas Smytheman, B.Sc., Emma Johnson, B.S., Valerie A. Reese, M.S., Susan L. Baldwin, Ph.D., Brittany Williams, B.Sc., Suhavi Kaur, B.Sc., Zhiyi Zhu, Ph.D.

**University of Rochester Medical Center, Rochester, NY, VTEU**

Angela R. Branche, MD, David Dobrzynski, MD, Ann R. Falsey, MD, Ian Shannon, MS, RN, Patrick Kingsley, Arthur Zemanek, RN, Sophia Wiltse, Erin Nowicki, Sharon Moorehead, Kari Steinmetz, CCRC, Doreen Francis, RN, CCRC, Cynthia MacDonald, RN, Jeanne Holden-Wiltse, MPH, MBA, Christopher Lane, Elizabeth Fladt, Kyle Richards, PharmD, Stephen Bean, PharmD, Nicole Dornbush, PharmD, Carol Cole, PharmD

**The Hope Clinic of Emory University (Hope), Atlanta, GA, VTEU**

Srilatha Edupuganti, M.D., M.P.H., Daniel Graciaa, M.D., Nadine Rouphael, M.D., M.Sc.,
Alexis Ahonen, M.S.N., Alicarmen Alvarez, R.N., Amy Anderson, R.N., Mary Atha, M.S.N., Sarah Bechnak, R.N., Mary Bower, R.N., Ellie Butler, Laura Clegg, R.N., Carla N. Cooke, P.A., Sharon Curate-Ingram, R.N., Renata Lynn Dennis, M.P.H., R.N., Francine Dyer, R.N., Srilatha Edupuganti, M.D., M.P.H., Tigisty Girmary, R.N., Rebecca Gonzalez, Pharm.D., Natalie Gray, Cassie Grimsley-Ackerley, M.D., M.Sc., Lauren Hewitt, L.P.N., Christopher

Huerta, Brandi Johnson, Colleen Kelley, M.D., M.P.H., Athena Koumanelis, Deborah Laryea, R.N., Cecilia Losada, Hollie Macenczak, R.N., Michelle Piane McCullough, Eileen Osinski, Bernadine Panganiban, Varun Phadke, M.D., Paulina A. Rebolledo, M.D., M.Sc., Nadine Rouphael, M.D., M.Sc., Erin Scherer, Ph.D., Jessica Traenkner, P.A., Kristen Unterberger, P.A., Jacob Usher, Michelle Wiles, R.N., T. Jean Winter, Jianguo Xu, R.Ph., Ph.D., Yongxian Xu

**Emory Children's Center (ECC), Atlanta, GA, VTEU**
Christina Rostad, M.D., Evan J. Anderson, M.D., Satoshi Kamidani, M.D., Amari Barrett, Julia Bartol, Leisa Bower, R.N., Jessica Bowman, R.N., Andres F. Camacho-Gonzalez, M.D., M.Sc., Victoria Curry, R.N., Theda Gibson, M.S., Felicia Glover, Hui-Mien Hsiao, M.S., Amberly Hunter, Laila Hussaini, M.P.H., Inara Jooma, Satoshi Kamidani, M.D., Peggy Kettle, R.N., Marcia Lewis, R.N., Wensheng Li, Cindy Lubbers, R.N., Lisa Macoy, R.N., M.S.N., Molly Morrison, Amy Muchinsky, Heather Nurse, R.N., Etza Peters, R.N., Susan Rogers, R.Ph., Christina A. Rostad, M.D., Amber Samuel, Maya Stagg, Kathy Stephens, R.N., M.S.N., Kathryn Zaks, M.S.

**University of Pittsburgh School of Medicine, Pittsburgh, PA, subsidiary to Vanderbilt University VTEU, Nashville, TN**
Judith M. Martin, M.D., Kumaravel Rajakumar, M.D., M.S., Gysella B. Muniz M.D., Sonika Bhatnagar, M.D., M.P.H., Alejandro Hoberman, MD, Timothy Shope MD, MPH, Nader Shaikh, MD, MPH, Spenser Kinsey BSN RN, Jennifer Opal RN, Melissa Andrasko RN, Shannon Mance MPH BSN RN, Lisa Vavro BSN RN, Lalicia Roman AS BS MS, Kimberly McMurtry PharmD, Emily Dougherty MPH, Kevin Lynch BS, Sabrina Catalano BS, Jamie Fries RN, MaryAnn Sieber RN, John F. Alcorn, Ph.D., Bo Zhai PhD

**University of Washington, Seattle, WA, VTEU**
Christine Johnston, M.D., M.P.H., Tara M. Babu, M.D., M.SCI., Anna Wald, M.D., M.P.H., Morissa Pertik, PA-C, T. Nui Pholsena, ARNP, Jina Taub, ARNP, Dana Varon, ARNP; Meredith Potochnic, PharmD; Matthew Dustrude, Kerry Laing, PhD, David M. Koelle, MD; Alyssa Braun, Anya Mathur, Dolly Singh, Jessica Heimonen, Jessica Moreno, Linsey McClellan, Maddie Humphreys, Ray Larsen, Taylor Krause, Mary Kirk, MPH, Matthew Seymour, MPH, Kirsten Hauge, MPH, Lawrence Hemingway, Christopher McClurkan, Max Krist, Victoria Campbell

**University of Texas Medical Branch (UTMB), League City, TX, subsidiary to BCM VTEU**
Richard E. Rupp, M.D., Megan Berman, M.D., Laura Porterfield, M.D., Amber Stanford, PA-C, Kristin Pollock, RN, Robert Cox, RN, Hala Ghoson, PharmD, Claire Marsh, BSN, RN, Amy McMahan, LVN, Esther Cox, Diane Barrett, MS;

**NYU VETU, New York, NY**
***New York University – Langone Vaccine Center, Manhattan, New York, NY, VTEU***
Mark J. Mulligan, MD, Mary Olson, NP, Marie Samanovic, PhD, Amber Cornelius, MS, Abdonnie Holder, James Wilson, Meron Tasissa, Hibah Khan, Sajjad Hussein, Doaa Ayoubi, Pharm.D., Ph.D., Mahnoor Ali

***NYU Langone Vaccine Center Research Clinic at Bellevue Hospital, New York, NY - subsidiary to NYU VTEU:***
Angelica Kottkamp, MD, Jennifer Dong, MD, Damian Inlall, Ellie Carmody, MD, Rebecca Boas, MD, Jennifer Knishinsky, MD, Reza Parungao, MD, Adam Schwartz, MD, Melinda Katz, MD, Natella Aronova, NP, Rita Mennuti,

NP, Athina Agbayani, RN, Zeeshan Iqbal, Julia Wagner, Venissala Wongchai, Tiffany Salcito, Leeja Abraham, Denise Dong, Lance Goodman, Nadia Tadros, Danhong Jiang, Anna Jacobs, Pharm.D., Alina Neganova, RN

***New York University- Long Island Vaccine Center, Mineola, NY, subsidiary to NYU VTEU***
Martín Bäcker, M.D., Steven E. Carsons, M.D., Kimberly Byrnes, R.N.CCRC, Diana Badillo, M.D., Asif Noor, M.D., Sigridh Muñoz-Gómez, M.D., Claudia De La Matta Rodriguez, M.D., Urmee Saha, M.D., Sajumon Joseph, F.N.P., Reny Jose, F.N.P, Catherine Tsiakaros, R.N., Sarah J. Pastolero, R.N., Anita Farhi, R.N., Maung Aung, Alicia Vasile, B.S., R.Ph., April Correll, R.Ph., Louis Ragolia, Ph.D., Christopher Hall, Thomas Palaia, Fiona T. Fitzgerald, Madalyn Saporito, Lavern Harvey.

**Cincinnati Children's Hospital Medical Center (CCHMC), Cincinnati, OH, VTEU**
Rebecca C. Brady, M.D.; Robert W. Frenck, M.D.; Paul W. Spearman, M.D.; Grant C. Paulsen, M.D.; Eleanor Widdice, M.D.; Felicia A. Scaggs Huang, M.D.; Michelle Dickey, A.P.R.N.; Kristen Buschle, A.P.R.N.; Susan Parker, R.N.; Stacy Ranz, L.P.N.; Tammy Lewis-McCauley, L.P.N.; Margery Huron, R.N.; Sally McCartney, R.N.; Jamie Kidd, R.N.; Jennifer Whitaker, R.N.; Kristie Price, Pharm.D.; Sarah Boland, R.Ph.; Jesse LePage, B.S.; Monica Malone McNeal, M.S.; Laura Pace, B.A.; Theresa Baker, M.S.; Robert Zoellner, B.S.; Mary Pat McKee, M.S.; Nicole Vollman, B.A., Kristie Price

**FHI360, Durham, NC**
Janet I. Archer M.Sc., Kuleni Abebe M.Sc. Heather Dickerson B.A., Sandra B. Brindis B.A., Linda McNeil MA PMP, Mary Briggs, Katlyn Hurst B.Sc.

**IDCRC Leadership and Administrative Team**
Kathleen M. Neuzil, M.D., David S. Stephens, M.D., Monica M. Farley M.D., Jeanne Marrazzo, M.D., Robert L. Atmar M.D., Jeffery Lennox, M.D., Sidnee Paschal Young, MALS; Barbara E. Walsh, Kayla Smith, MPH; and Bridget A. Wynn

**IDCRC Statistical and Data Science Unit (SDSU), Vaccine and Infectious Disease Division, Fred Hutchinson Cancer Research Center, Seattle, WA:**
Elizabeth R. Brown, Sc.D., Clara Dominguez Islas, Ph.D.

**Statistical Center for HIV/AIDS Research and Prevention (SCHARP), Fred Hutchinson Cancer Research Center, Seattle, WA:**
Jillian Zemanek, M.P.H., Daniel Szydlo, M.S., Rahul PaulChoudhury, M.S., Brian Ingersoll, Wen-Min (Wendy) Hou, M.P.H., B.S.N, C.C.R.P., Kelly Maddox, Chloe Waters, Lauren Young, M.P.H., Karan A. Shah, M.S., Anisa C. Gravelle, M.S., Craig N Chin, Drew Edwards, Rudie Desravines, M.S., Mark Trumbull, M.S., Srikanth Nooney, Udhab Adhikari, Josh Larkin, Jackie Benson, Christine Thompson, Jean-Paul Pease.

**IDCRC Laboratory Operations Unit (LOU):** Christine M. Posavad, PhD; John Hural, PhD; Michael Stirewalt; Megan Meagher

**Division of Microbiology and Infectious Diseases, National Institute of Allergy and Infectious Diseases, National Institutes of Health, Bethesda, MD.**
Marina Lee, Ph.D., Mohamed Elsafy, M.D., Rhonda Pikaart-Tautges, B.S. Janice Arega, M.S., Binh Hoang, R. Ph., Dan Curtin, Olivia Sparer, B.A., Ranjodh Gill M.P.H, Hyung Koo, B.S.N., Elisa Sindall, B.S.N., Sonja Crandon, B.S.N., Seema Nayak, M.D., Diane J. Post, Ph. D., Paul C Roberts, Ph.D., John Beigel, M.D.

**Thermo Fisher Scientific, Germantown, MD.**
Jim Dunn

**Duke University Laboratory – Durham, NC**
David Montefiori, Ph.D., Amanda Eaton, Ph.D.

### Supplemental Methods

<u>Pseudovirus Neutralization Immunogenicity Method Description</u>

The Duke NAb Laboratory for HIV and COVID-19 Vaccine Research and Development (Duke NAb Lab; PI: Dr. David Montefiori) assessed the magnitude and cross-variant SARS-CoV-2 neutralizing antibody responses in the DMID Protocol #21-0012 by using a fully validated assay in an environment that operates in compliance with Good Clinical Laboratory Practices (GCLP). The endpoint reported here is the SARS-CoV-2-specific neutralizing activity of antibodies in serum. Neutralizing antibodies were assessed with Spike-pseudotyped viruses in 293T/ACE2 cells as a function of reductions in firefly luciferase reporter activity.[1] Assays were performed on all samples with Spike-pseudotyped SARS-CoV-2 D614G and Omicron. Results are reported as Inhibitory Dilution 50 (ID50); these values represent the serum dilution that reduces relative luminescence units (RLU) by 50% relative to the RLU in the virus control wells after subtraction of background RLU. This study was completed under the oversight of the Quality Assurance Unit for Duke Vaccine Immunogenicity Programs (QADVIP).

The Spike mutations listed below were in the pseudovirus variants used in the assay:

| **Variant** | **Spike mutations** |
|---|---|
| D614G | D614G |
| Omicron | A67V, Δ69-70, T95I, G142D, Δ143-145, Δ211, L212I, +214EPE, G339D, S371L, S373P, S375F, K417N, N440K, G446S, S477N, T478K, E484A, Q493R, G496S, Q498R, N501Y, Y505H, T547K, D614G, H655Y, N679K, P681H, N764K, D796Y, N856K, Q954H, N969K, L981F |

Statistical Methods for Sample Selection

Twenty participants from each enrollment group were randomly selected for Omicron testing, stratified by age group so that 10 of the participants selected in each enrollment group were 18-55 years of age and 10 were 56 years of age or older. Only participants with sufficient serum collected to enable testing were considered for selection.

**Supplemental Table 1.** SARS-CoV-2 Pseudovirion Neutralization Antibody Titers (ID50) to Pseudovirus D614G[1], by EUA Group, Booster, and Timepoint

| | Group 2E [Dosed Moderna, Boost Moderna 100-mcg] (N=20) | Group 13E [Dosed Moderna, Boost Moderna 1273 50-mcg] (N=20) | Group 4E [Dosed Janssen, Boost Janssen] (N=20) | Group 6E [Dosed Pfizer/BioNtech, Boost Janssen] (N=20) | Group 7E [Dosed Janssen, Boost Pfizer] (N=20) | Group 9E [Dosed Pfizer/BioNtech, Boost Pfizer] (N=20) |
|---|---|---|---|---|---|---|
| **Day 1 Visit (Pre-boost)** | | | | | | |
| N (non-missing) | 20 | 20 | 20 | 20 | 20 | 20 |
| Positive Response(%)[2] | 20 (100.0%) | 20 (100.0%) | 17 ( 85.0%) | 19 ( 95.0%) | 18 ( 90.0%) | 19 ( 95.0%) |
| Median ($P_{25}$, $P_{75}$) | 357.54 (192.38-705.12) | 135.14 (69.72-242.29) | 32.28 (18.01-65.42) | 89.14 (33.24-143.07) | 41.46 (21.16-63.34) | 89.48 (41.06-206.74) |
| Minimum - Maximum | 99.29-1420.81 | 13.02-1127.65 | 5.00-3447.06 | 5.00-604.29 | 5.00-377.04 | 5.00-608.99 |
| Geometric Mean (95% CI) | 342.72 (233.42-503.19) | 129.54 (77.83-215.60) | 35.28 (17.89-69.61) | 71.46 (41.21-123.90) | 40.89 (23.36-71.56) | 94.25 (53.37-166.43) |
| **Day 29 Visit (28 days post-boost)** | | | | | | |
| N (non-missing) | 20 | 20 | 20 | 20 | 20 | 20 |
| Positive Response(%)[2] | 20 (100.0%) | 20 (100.0%) | 20 (100.0%) | 20 (100.0%) | 20 (100.0%) | 20 (100.0%) |
| Median ($P_{25}$, $P_{75}$) | 2816.43 (1388.91-4214.12) | 4322.97 (3741.11-5550.53) | 134.01 (71.18-177.76) | 1187.64 (630.96-2027.19) | 897.75 (713.86-1728.09) | 981.78 (777.52-2576.32) |
| Minimum - Maximum | 813.55-14150.83 | 590.89-14881.03 | 25.21-2460.53 | 146.04-6161.39 | 386.53-2910.15 | 424.96-5603.55 |
| Geometric Mean (95% CI) | 2708.49 (1878.59-3905.02) | 3941.98 (2772.63-5604.51) | 127.58 (76.51-212.73) | 1017.78 (633.81-1634.35) | 1044.90 (808.42-1350.54) | 1315.53 (932.46-1855.97) |
| N* (non-missing pre- and post-boost) | 20 | 20 | 20 | 20 | 20 | 20 |
| Participants with ≥ 2-fold rise[3], 95% CI | 100.0% (83.2%-100.0%) | 100.0% (83.2%-100.0%) | 60.0% (36.1%-80.9%) | 100.0% (83.2%-100.0%) | 100.0% (83.2%-100.0%) | 95.0% (75.1%-99.9%) |
| Participants with ≥ 4-fold rise[3], 95% CI | 80.0% (56.3%-94.3%) | 100.0% (83.2%-100.0%) | 50.0% (27.2%-72.8%) | 85.0% (62.1%-96.8%) | 90.0% (68.3%-98.8%) | 90.0% (68.3%-98.8%) |
| Geometric Mean Fold Rise[3], 95% CI | 7.90 (5.54-11.27) | 30.43 (18.44-50.23) | 3.62 (2.02-6.48) | 14.24 (8.26-24.57) | 25.55 (14.43-45.26) | 13.96 (7.67-25.39) |

[1] Values below the lower limit of detection (LLOD = 10) were assigned the value of 5 (LLOD/2).

[2] Positive response is defined as an ID50 titer above the LLOD (ID50 ≥10).

[3] Relative to pre-vaccination (Day 1 Visit) levels, among participants with non-missing observations at both pre- and post-boost timepoints.

**Supplemental Table 2.** SARS-CoV-2 Pseudovirion Neutralization Antibody Titers (ID50) to Pseudovirus D614G[1], by EUA Group, Booster, Timepoint, and Age

| | Group 2E [Dosed Moderna, Boost Moderna 100-mcg] Age 18-55 yo (N=10) | Group 2E [Dosed Moderna, Boost Moderna 100-mcg] Age ≥56 yo (N=10) | Group 13E [Dosed Moderna, Boost Moderna 1273 50-mcg] Age 18-55 yo (N=10) | Group 13E [Dosed Moderna, Boost Moderna 1273 50-mcg] Age ≥56 yo (N=10) | Group 4E [Dosed Janssen, Boost Janssen] Age 18-55 yo (N=10) | Group 4E [Dosed Janssen, Boost Janssen] Age ≥56 yo (N=10) |
|---|---|---|---|---|---|---|
| **Day 1 Visit (Pre-boost)** | | | | | | |
| N (non-missing) | 10 | 10 | 10 | 10 | 10 | 10 |
| Positive Response(%)[2] | 10 (100.0%) | 10 (100.0%) | 10 (100.0%) | 10 (100.0%) | 9 ( 90.0%) | 8 ( 80.0%) |
| Median ($P_{25}$, $P_{75}$) | 357.54 (193.39-592.40) | 403.19 (191.36-715.45) | 219.20 (121.05-263.35) | 85.61 (32.55-149.24) | 34.09 (26.94-87.46) | 27.50 (15.12-57.26) |
| Minimum - Maximum | 104.33-771.93 | 99.29-1420.81 | 68.53-1127.65 | 13.02-253.06 | 5.00-3447.06 | 5.00-95.77 |
| Geometric Mean (95% CI) | 307.28 (183.25-515.27) | 382.24 (195.91-745.80) | 230.79 (122.81-433.71) | 72.71 (35.78-147.75) | 50.96 (14.39-180.50) | 24.43 (11.84-50.39) |
| **Day 29 Visit (28 days post-boost)** | | | | | | |
| N (non-missing) | 10 | 10 | 10 | 10 | 10 | 10 |
| Positive Response(%)[2] | 10 (100.0%) | 10 (100.0%) | 10 (100.0%) | 10 (100.0%) | 10 (100.0%) | 10 (100.0%) |
| Median ($P_{25}$, $P_{75}$) | 2458.46 (1607.99-3281.39) | 3357.07 (1286.86-7479.37) | 4064.94 (3736.80-5126.41) | 4764.57 (4131.29-6660.04) | 153.37 (105.40-198.18) | 119.07 (58.94-153.47) |
| Minimum - Maximum | 813.55-4822.79 | 1068.30-14150.83 | 1079.06-8357.64 | 590.89-14881.03 | 28.07-2460.53 | 25.21-552.98 |
| Geometric Mean (95% CI) | 2278.58 (1506.90-3445.44) | 3219.51 (1639.65-6321.61) | 3698.68 (2437.90-5611.50) | 4201.29 (2176.80-8108.59) | 163.75 (67.65-396.34) | 99.40 (51.32-192.55) |
| N* (non-missing pre- and post-boost) | 10 | 10 | 10 | 10 | 10 | 10 |
| Participants with ≥ 2-fold rise[3], 95% CI | 100.0% (69.2%-100.0%) | 100.0% (69.2%-100.0%) | 100.0% (69.2%-100.0%) | 100.0% (69.2%-100.0%) | 70.0% (34.8%-93.3%) | 50.0% (18.7%-81.3%) |
| Participants with ≥ 4-fold rise[3], 95% CI | 80.0% (44.4%-97.5%) | 80.0% (44.4%-97.5%) | 100.0% (69.2%-100.0%) | 100.0% (69.2%-100.0%) | 50.0% (18.7%-81.3%) | 50.0% (18.7%-81.3%) |
| Geometric Mean Fold Rise[3], 95% CI | 7.42 (4.65-11.83) | 8.42 (4.48-15.84) | 16.03 (8.43-30.48) | 57.78 (31.76-105.12) | 3.21 (1.41-7.32) | 4.07 (1.51-10.98) |

| | Group 6E [Dosed Pfizer/BioNTech, Boost Janssen] Age 18-55 yo (N=10) | Group 6E [Dosed Pfizer/BioNTech, Boost Janssen] Age ≥56 yo (N=10) | Group 7E [Dosed Janssen, Boost Pfizer] Age 18-55 yo (N=10) | Group 7E [Dosed Janssen, Boost Pfizer] Age ≥56 yo (N=10) | Group 9E [Dosed Pfizer/BioNTech, Boost Pfizer] Age 18-55 yo (N=10) | Group 9E [Dosed Pfizer/BioNTech, Boost Pfizer] Age ≥56 yo (N=10) |
|---|---|---|---|---|---|---|
| **Day 1 Visit (Pre-boost)** | | | | | | |
| N (non-missing) | 10 | 10 | 10 | 10 | 10 | 10 |
| Positive Response(%)[2] | 10 (100.0%) | 9 ( 90.0%) | 9 ( 90.0%) | 9 ( 90.0%) | 10 (100.0%) | 9 ( 90.0%) |
| Median ($P_{25}$, $P_{75}$) | 95.61 (43.57-162.94) | 66.17 (32.42-123.21) | 41.17 (21.49-198.05) | 41.46 (20.13-56.81) | 188.23 (87.13-436.03) | 74.86 (28.18-91.82) |
| Minimum - Maximum | 14.79-526.28 | 5.00-604.29 | 5.00-377.04 | 5.00-192.31 | 32.61-608.99 | 5.00-174.59 |
| Geometric Mean (95% CI) | 84.28 (39.56-179.54) | 60.59 (23.57-155.74) | 51.09 (19.20-135.93) | 32.73 (15.79-67.82) | 168.71 (76.43-372.44) | 52.65 (24.55-112.92) |
| **Day 29 Visit (28 days post-boost)** | | | | | | |
| N (non-missing) | 10 | 10 | 10 | 10 | 10 | 10 |
| Positive Response(%)[2] | 10 (100.0%) | 10 (100.0%) | 10 (100.0%) | 10 (100.0%) | 10 (100.0%) | 10 (100.0%) |
| Median ($P_{25}$, $P_{75}$) | 1630.35 (699.99-2718.12) | 997.26 (334.54-1475.96) | 1056.50 (849.81-1802.61) | 713.86 (532.02-1706.98) | 1000.04 (770.06-2881.47) | 897.66 (784.98-2271.16) |
| Minimum - Maximum | 146.04-6161.39 | 161.56-3260.65 | 804.76-2910.15 | 386.53-1985.60 | 424.96-5603.55 | 631.66-3108.24 |
| Geometric Mean (95% CI) | 1309.31 (617.02-2778.30) | 791.16 (399.11-1568.32) | 1238.02 (884.96-1731.94) | 881.90 (577.46-1346.84) | 1414.08 (763.14-2620.25) | 1223.85 (784.42-1909.43) |
| N* (non-missing pre- and post-boost) | 10 | 10 | 10 | 10 | 10 | 10 |
| Participants with ≥ 2-fold rise[3], 95% CI | 100.0% (69.2%-100.0%) | 100.0% (69.2%-100.0%) | 100.0% (69.2%-100.0%) | 100.0% (69.2%-100.0%) | 90.0% (55.5%-99.7%) | 100.0% (69.2%-100.0%) |
| Participants with ≥ 4-fold rise[3], 95% CI | 100.0% (69.2%-100.0%) | 70.0% (34.8%-93.3%) | 90.0% (55.5%-99.7%) | 90.0% (55.5%-99.7%) | 80.0% (44.4%-97.5%) | 100.0% (69.2%-100.0%) |
| Geometric Mean Fold Rise[3], 95% CI | 15.54 (9.17-26.32) | 13.06 (4.41-38.71) | 24.23 (8.49-69.16) | 26.95 (13.21-54.98) | 8.38 (3.55-19.76) | 23.24 (9.86-54.78) |

[1] Values below the lower limit of detection (LLOD = 10) were assigned the value of 5 (LLOD/2).
[2] Positive response is defined as an ID50 titer above the LLOD (ID50 ≥10).
[3] Relative to pre-vaccination (Day 1 Visit) levels, among participants with non-missing observations at both pre- and post-boost timepoints.

**Supplemental Table 3.** SARS-CoV-2 Pseudovirion Neutralization Antibody Titers (ID50) to Pseudovirus Omicron[1], by EUA Group, Booster, and Timepoint

| | Group 2E [Dosed Moderna, Boost Moderna 100-mcg] (N=20) | Group 13E [Dosed Moderna, Boost Moderna 1273 50-mcg] (N=20) | Group 4E [Dosed Janssen, Boost Janssen] (N=20) | Group 6E [Dosed Pfizer/BioNtech, Boost Janssen] (N=20) | Group 7E [Dosed Janssen, Boost Pfizer] (N=20) | Group 9E [Dosed Pfizer/BioNtech, Boost Pfizer] (N=20) |
|---|---|---|---|---|---|---|
| **Day 1 Visit (Pre-boost)** | | | | | | |
| N (non-missing) | 20 | 20 | 20 | 20 | 20 | 19 |
| Positive Response(%)[2] | 18 ( 90.0%) | 17 ( 85.0%) | 4 ( 20.0%) | 10 ( 50.0%) | 5 ( 25.0%) | 15 ( 78.9%) |
| Median ($P_{25}$, $P_{75}$) | 25.32 (15.48-32.02) | 17.29 (12.45-25.79) | 5.00 (5.00-5.00) | 8.28 (5.00-35.73) | 5.00 (5.00-8.49) | 19.57 (10.34-31.87) |
| Minimum - Maximum | 5.00-140.61 | 5.00-100.95 | 5.00-356.01 | 5.00-110.81 | 5.00-101.86 | 5.00-52.16 |
| Geometric Mean (95% CI) | 24.53 (16.48-36.51) | 18.55 (12.33-27.88) | 7.16 (4.51-11.38) | 12.75 (7.64-21.28) | 7.64 (5.15-11.34) | 17.31 (11.88-25.23) |
| GM Fold Decrease Relative to D614G[3], 95% CI | 13.97 (9.53-20.49) | 6.98 (5.27-9.26) | 6.53 (4.34-9.82) | 6.13 (4.58-8.22) | 6.45 (4.20-9.90) | 6.03 (3.38-10.77) |
| **Day 29 Visit (28 days post-boost)** | | | | | | |
| N (non-missing) | 20 | 20 | 20 | 20 | 20 | 20 |
| Positive Response(%)[2] | 20 (100.0%) | 20 (100.0%) | 20 (100.0%) | 20 (100.0%) | 20 (100.0%) | 20 (100.0%) |
| Median ($P_{25}$, $P_{75}$) | 393.11 (197.64-833.88) | 699.95 (341.80-1624.85) | 33.14 (21.96-69.96) | 373.89 (131.32-1170.62) | 227.34 (150.95-701.52) | 530.04 (311.73-971.22) |
| Minimum - Maximum | 60.34-1232.53 | 42.41-6974.31 | 14.89-879.05 | 36.63-3140.44 | 46.45-1058.09 | 182.63-2319.52 |
| Geometric Mean (95% CI) | 361.70 (233.06-561.35) | 712.08 (404.79-1252.64) | 45.04 (27.01-75.12) | 348.03 (188.24-643.47) | 266.18 (173.50-408.37) | 571.05 (406.82-801.59) |
| GM Fold Decrease Relative to D614G[3], 95% CI | 7.49 (5.58-10.04) | 5.54 (4.18-7.34) | 2.83 (2.28-3.51) | 2.92 (2.10-4.07) | 3.93 (2.65-5.82) | 2.30 (1.74-3.05) |
| N* (non-missing pre- and post-boost) | 20 | 20 | 20 | 20 | 20 | 19 |
| Participants with ≥ 2-fold rise[4], 95% CI | 100.0% (83.2%-100.0%) | 100.0% (83.2%-100.0%) | 100.0% (83.2%-100.0%) | 100.0% (83.2%-100.0%) | 100.0% (83.2%-100.0%) | 100.0% (82.4%-100.0%) |
| Participants with ≥ 4-fold rise[4], 95% CI | 90.0% (68.3%-98.8%) | 100.0% (83.2%-100.0%) | 65.0% (40.8%-84.6%) | 100.0% (83.2%-100.0%) | 95.0% (75.1%-99.9%) | 100.0% (82.4%-100.0%) |
| Geometric Mean Fold Rise[4], 95% CI | 14.75 (10.13-21.46) | 38.40 (24.39-60.45) | 6.29 (4.12-9.59) | 27.29 (17.59-42.34) | 34.84 (22.37-54.25) | 30.64 (20.04-46.84) |

[1] Values below the lower limit of detection (LLOD = 10) were assigned the value of 5 (LLOD/2).

[2] Positive response is defined as an ID50 titer above the LLOD (ID50 ≥10).

[3] Relative to pre-vaccination (Day 1 Visit) levels, among participants with non-missing observations at both pre- and post-boost timepoints.

**Supplemental Table 4.** SARS-CoV-2 Pseudovirion Neutralization Antibody Titers (ID50) to Pseudovirus B.1.1.529[1], by EUA Group, Booster, Timepoint, and Age

| | Group 2E [Dosed Moderna, Boost Moderna 100-mcg] Age 18-55 yo (N=10) | Group 2E [Dosed Moderna, Boost Moderna 100-mcg] Age ≥56 yo (N=10) | Group 13E [Dosed Moderna, Boost Moderna 1273 50-mcg] Age 18-55 yo (N=10) | Group 13E [Dosed Moderna, Boost Moderna 1273 50-mcg] Age ≥56 yo (N=10) | Group 4E [Dosed Janssen, Boost Janssen] Age 18-55 yo (N=10) | Group 4E [Dosed Janssen, Boost Janssen] Age ≥56 yo (N=10) |
|---|---|---|---|---|---|---|
| **Day 1 Visit (Pre-boost)** | | | | | | |
| N (non-missing) | 10 | 10 | 10 | 10 | 10 | 10 |
| Positive Response(%)[2] | 8 ( 80.0%) | 10 (100.0%) | 9 ( 90.0%) | 8 ( 80.0%) | 2 ( 20.0%) | 2 ( 20.0%) |
| Median ($P_{25}$, $P_{75}$) | 25.32 (19.32-28.67) | 25.90 (15.36-44.94) | 19.74 (14.41-88.04) | 15.73 (10.02-18.70) | 5.00 (5.00-5.00) | 5.00 (5.00-5.00) |
| Minimum - Maximum | 5.00-129.76 | 12.07-140.61 | 5.00-100.95 | 5.00-30.34 | 5.00-356.01 | 5.00-13.28 |
| Geometric Mean (95% CI) | 21.51 (11.05-41.87) | 27.97 (15.93-49.13) | 25.55 (12.40-52.66) | 13.46 (8.80-20.59) | 8.64 (3.26-22.85) | 5.94 (4.57-7.73) |
| GM Fold Decrease Relative to D614G[3], 95% CI | 14.28 (9.47-21.55) | 13.66 (6.49-28.75) | 9.03 (6.24-13.08) | 5.40 (3.53-8.26) | 7.19 (3.65-14.16) | 5.86 (3.19-10.76) |
| **Day 29 Visit (28 days post-boost)** | | | | | | |
| N (non-missing) | 10 | 10 | 10 | 10 | 10 | 10 |
| Positive Response(%)[2] | 10 (100.0%) | 10 (100.0%) | 10 (100.0%) | 10 (100.0%) | 10 (100.0%) | 10 (100.0%) |
| Median ($P_{25}$, $P_{75}$) | 393.11 (145.82-716.43) | 430.50 (220.93-933.84) | 784.06 (434.55-1231.87) | 680.27 (249.05-2196.18) | 642.87 (292.26-991.16) | 530.04 (331.19-950.04) |
| Minimum - Maximum | 60.34-1105.87 | 62.93-1232.53 | 143.08-3547.49 | 42.41-6974.31 | 182.63-1829.91 | 248.43-2319.52 |
| Geometric Mean (95% CI) | 339.92 (172.81-668.60) | 384.89 (191.03-775.49) | 706.71 (361.43-1381.82) | 717.49 (248.56-2071.13) | 562.17 (325.64-970.48) | 580.08 (345.41-974.20) |
| GM Fold Decrease Relative to D614G[3], 95% CI | 6.70 (4.17-10.76) | 8.36 (5.43-12.88) | 5.23 (3.78-7.25) | 5.86 (3.44-9.96) | 2.52 (1.74-3.64) | 2.11 (1.29-3.44) |
| N* (non-missing pre- and post-boost) | 10 | 10 | 10 | 10 | 10 | 9 |
| Participants with ≥ 2-fold rise[4], 95% CI | 100.0% (69.2%-100.0%) | 100.0% (69.2%-100.0%) | 100.0% (69.2%-100.0%) | 100.0% (69.2%-100.0%) | 100.0% (69.2%-100.0%) | 100.0% (66.4%-100.0%) |
| Participants with ≥4-fold rise[4], 95% CI | 100.0% (69.2%-100.0%) | 80.0% (44.4%-97.5%) | 100.0% (69.2%-100.0%) | 100.0% (69.2%-100.0%) | 100.0% (69.2%-100.0%) | 100.0% (66.4%-100.0%) |
| Geometric Mean Fold Rise[4], 95% CI | 15.80 (10.11-24.70) | 13.76 (6.83-27.70) | 27.66 (15.52-49.28) | 53.30 (25.22-112.68) | 29.56 (18.21-47.98) | 31.88 (13.61-74.69) |

| | Group 6E [Dosed Pfizer/BioNTech, Boost Janssen] Age 18-55 yo (N=10) | Group 6E [Dosed Pfizer/BioNTech, Boost Janssen] Age ≥56 yo (N=10) | Group 7E [Dosed Janssen, Boost Pfizer] Age 18-55 yo (N=10) | Group 7E [Dosed Janssen, Boost Pfizer] Age ≥56 yo (N=10) | Group 9E [Dosed Pfizer/BioNTech, Boost Pfizer] Age 18-55 yo (N=10) | Group 9E [Dosed Pfizer/BioNTech, Boost Pfizer] Age ≥56 yo (N=10) |
|---|---|---|---|---|---|---|
| **Day 1 Visit (Pre-boost)** | | | | | | |
| N (non-missing) | 10 | 10 | 10 | 10 | 10 | 9 |
| Positive Response (%)[1] | 5 ( 50.0%) | 5 ( 50.0%) | 3 ( 30.0%) | 2 ( 20.0%) | 8 ( 80.0%) | 7 ( 77.8%) |
| Median ($P_{25}$, $P_{75}$) | 9.25 (5.00-37.43) | 8.28 (5.00-22.16) | 5.00 (5.00-15.85) | 5.00 (5.00-5.00) | 25.35 (12.69-33.05) | 16.42 (10.34-31.15) |
| Minimum - Maximum | 5.00-110.81 | 5.00-75.44 | 5.00-101.86 | 5.00-31.30 | 5.00-45.01 | 5.00-52.16 |
| Geometric Mean (95% CI) | 14.42 (5.98-34.76) | 11.28 (5.56-22.89) | 8.91 (4.24-18.68) | 6.56 (4.22-10.17) | 19.02 (10.83-33.39) | 15.60 (8.38-29.05) |
| GM Fold Decrease Relative to D614G[2], 95% CI | 5.85 (3.67-9.32) | 6.47 (4.12-10.15) | 6.97 (3.73-13.02) | 5.97 (2.87-12.41) | 8.87 (3.97-19.80) | 3.72 (1.49-9.31) |
| **Day 29 Visit (28 days post-boost)** | | | | | | |
| N (non-missing) | 10 | 10 | 10 | 10 | 10 | 10 |
| Positive Response (%)[1] | 10 (100.0%) | 10 (100.0%) | 10 (100.0%) | 10 (100.0%) | 10 (100.0%) | 10 (100.0%) |
| Median ($P_{25}$, $P_{75}$) | 724.90 (177.81-1168.97) | 150.14 (98.87-1172.26) | 288.37 (215.65-688.28) | 212.04 (131.48-843.87) | 642.87 (292.26-991.16) | 530.04 (331.19-950.04) |
| Minimum - Maximum | 47.97-3140.44 | 36.63-1276.45 | 46.45-842.88 | 72.77-1058.09 | 182.63-1829.91 | 248.43-2319.52 |
| Geometric Mean (95% CI) | 517.30 (205.81-1300.19) | 234.15 (93.90-583.87) | 281.88 (148.42-535.35) | 251.36 (124.97-505.56) | 562.17 (325.64-970.48) | 580.08 (345.41-974.20) |
| GM Fold Decrease Relative to D614G[2], 95% CI | 2.53 (1.73-3.70) | 3.38 (1.84-6.22) | 4.39 (2.54-7.59) | 3.51 (1.79-6.87) | 2.52 (1.74-3.64) | 2.11 (1.29-3.44) |
| N* (non-missing pre- and post-boost) | 10 | 10 | 10 | 10 | 10 | 9 |
| Participants with ≥ 2-fold rise[3], 95% CI | 100.0% (69.2%-100.0%) | 100.0% (69.2%-100.0%) | 100.0% (69.2%-100.0%) | 100.0% (69.2%-100.0%) | 100.0% (69.2%-100.0%) | 100.0% (66.4%-100.0%) |
| Participants with ≥4-fold rise[3], 95% CI | 100.0% (69.2%-100.0%) | 100.0% (69.2%-100.0%) | 90.0% (55.5%-99.7%) | 100.0% (69.2%-100.0%) | 100.0% (69.2%-100.0%) | 100.0% (66.4%-100.0%) |
| Geometric Mean Fold Rise[3], 95% CI | 35.88 (19.83-64.95) | 20.75 (10.13-42.52) | 31.65 (13.94-71.87) | 38.35 (22.50-65.35) | 29.56 (18.21-47.98) | 31.88 (13.61-74.69) |

[1] Values below the lower limit of detection (LLOD = 10) are assigned the value of 5 (LLOD/2).

[2] Relative to D614G titer at same visit, among participants with positive responses to both.

[3] Relative to pre-vaccination (Day 1 Visit) levels, among participants with non-missing observations at both pre- and post-boost timepoints.

**Supplemental Table 5.** Study Design by Stage and Group

| Group | Sample Size* | EUA Dosing Scheme | Interval (weeks) | Delayed Booster Vaccination | Strategy Tested |
|---|---|---|---|---|---|
| 2E | 50: 25 18-55 years, 25 ≥56 years | Previously dosed Moderna – mRNA-1273 | ≥12 | Moderna- mRNA-1273 | Control - Same Strain & platform |
| 4E | 50: 25 18-55 years, 25 ≥56 years | Previously dosed Janssen – Ad26.COV2-S | ≥12 | Janssen – Ad26.COV2.S | Control - Same Strain & platform |
| 6E | 50: 25 18-55 years, 25 ≥56 years | Previously dosed Pfizer/BioNTech –BNT162b2 | ≥12 | Janssen – Ad26.COV2.S | Same Strain Heterologous platform |
| 7E | 50: 25 18-55 years, 25 ≥56 years | Previously dosed Janssen – Ad26.COV2-S | ≥12 | Pfizer/BioNTech – BNT162b2 | Same Strain Heterologous platform |
| 9E | 50: 25 18-55 years, 25 ≥56 years | Previously dosed Pfizer/BioNTech –BNT162b2 | ≥12 | Pfizer/BioNTech – BNT162b2 | Control - Same Strain & platform |
| 13E | 50: 25 18-55 years, 25 ≥56 years | Previously dosed Moderna – mRNA-1273 | ≥12 | Moderna- mRNA-1273 | Control - Same Strain & platform |
